# Hypocalcemia and Acute Pulmonary Embolism Hospitalizations in the United States: Highlights from the propensity matched 2017 Nationwide Inpatient Sample

**DOI:** 10.1101/2021.07.22.21260999

**Authors:** Mukunthan Murthi, Hafeez Shaka, Zain El-amir, Sujitha Velagapudi, Abdul Jamil, Farah Wani, Ramtej Atluri, Akshay Kumar, Asim Kichloo

**Affiliations:** John H Stroger Hospital of Cook County; Central Michigan University College of Medicine, Department of Internal Medicine, Saginaw MI, USA; Samaritan Medical Center, Department of Internal Medicine, Watertown, NY, USA; Department of Cardiothoracic Surgery, University of Pittsburgh Medical Centre

**Keywords:** Hypocalcemia, Pulmonary embolism, mortality

## Abstract

Acute pulmonary embolism (PE) is a common cause for hospitalization associated with significant mortality and morbidity. Disorders of calcium metabolism are a frequently encountered medical problem. The effect of hypocalcemia is not well defined on the outcomes of patients with PE. We aimed to identify the prognostic value of hypocalcemia in hospitalized PE patients utilizing the 2017 Nationwide Inpatient Sample (NIS). In this retrospective study, we selected patients with a primary diagnosis of Acute PE using ICD 10 codes. They were further stratified based on the presence of hypocalcemia. We primarily aimed to compare in-hospital mortality for PE patients with and without hypocalcemia. In the 2017 NIS, 187,989 patients had a principal diagnosis of acute PE. Among the above study group, 1565(0.8%)had an additional diagnosis of hypocalcemia. 12.4% of PE patients with hypocalcemia died in the hospital in comparison to 2.95% without hypocalcemia. On multivariate regression analysis, PE and hypocalcemia patients had 4 times higher odds (aOR-4.03, 95% CI 2.78-5.84, p<0.001) of in-hospital mortality compared to those with only PE. We observed a similarly high odds of mortality(aOR=4.4) on 1:1 propensity-matched analysis. The incidence of acute kidney injury (aOR=2.62, CI 1.95-3.52, p<0.001), acute respiratory failure (a0R=1.84, CI 1.42-2.38, p<0.001), sepsis (aOR=4.99, CI 3.08-8.11, p<0.001) and arrhythmias (aOR=2.63, CI 1.99-3.48, p<0.001) were also higher for PE patients with hypocalcemia. Thus, PE patients with hypocalcemia have higher in-hospital complications and mortality than those without hypocalcemia.

## INTRODUCTION

Pulmonary embolism (PE) is a common medical condition associated with significant morbidity and mortality. Although the exact incidence of PE is not known, approximately 900,000 persons are affected by DVT/PE per year in the United States. It is associated with significant mortality and morbidity, with sudden death being the first symptom in 25% of the patients (1). Calcium plays a crucial role in multiple steps of the coagulation cascade including platelet adhesion, protease complex assembly and enzyme activation (2). Hypocalcemia is associated with the progression of and poorer prognosis of numerous cardiopulmonary diseases (3, 4). Calcium also is a part of the coagulation factor IV and thus is involved in the coagulation process (5).

Various factors are known to affect the outcomes of PE. One such factor being hypocalcemia. Previously, it was suspected to be a potential reason for PE by inducing lower extremity spasms (6). Validated prognostic scores like the Pulmonary Embolism Severity Index are currently widely used to determine mortality and morbidity in patients with PE(5). However, there is no evidence defining the effect of hypocalcemia on the outcomes of patients hospitalized with PE. We aimed to identify the prognostic value of hypocalcemia in hospitalized PE patients utilizing the 2017 Nationwide Inpatient Sample.

## METHODS

We utilized the Nationwide Inpatient Sample (NIS) database of 2017 to perform this retrospective study. The NIS is the largest publicly available all-payer inpatient healthcare database. It approximates a 20% stratified sample of discharges from community hospitals and provides an estimate of inpatient healthcare utilization, charges, and outcomes. It estimates more than 35 million hospitalizations every year. The data available in NIS includes primary and secondary diagnosis during hospitalization, demographic characteristics, inpatient procedures, hospital charges, and overall outcome. We utilized ICD-10 codes to identify the diagnosis. Our study did not require Institutional review board (IRB) approval since NIS data is de-identified. We selected patients with a primary diagnosis of Acute Pulmonary embolism (PE) using the ICD-10 code. They were further stratified based on the presence of hypocalcemia using ICD-10 codes.

We primarily aimed to compare in-hospital mortality for PE patients with and without hypocalcemia. Secondary outcomes included the effect of hypocalcemia on the incidence of acute kidney injury (AKI), Acute respiratory failure (ARF), arrhythmias, sepsis, length of hospital stay (LOS) and total hospital charges (THC).

We used STATA® (StataCorp, College Station, TX) version 16.1 to perform statistical analysis. Healthcare Cost and Utilization Project (HCUP) supplies year-based discharge weights to calculate weighted national estimates. Proportions were compared using Fisher’s exact test. Independent sample t-test was used to compare the mean of continuous data. Univariate regression analysis was used to calculate the unadjusted odds ratio (OR) for each outcome. Based on the univariate screen significance(p<0.2), we selected variables to perform a multivariate logistic regression analysis to adjust for potential confounders. The multivariate regression model also included variables that were considered essential based on a literary review. Logistic regression was used for binary outcomes and linear regression for continuous outcomes. All p values were two-tailed and we used a threshold of 0.05 to determine significance. Using the Charlson comorbidity index, we categorized patients into four groups based on 0, 1, 2, and ≥3 comorbidities

We used propensity scores to match patients with PE who had hypocalcemia to those who did not. The resultant model had a 1:1 match of similar groups. The propensity-matched cohort of patients was then used to compare the mortality.

## RESULTS

Among all patient encounters in the 2017 NIS, 187,989 had a principal diagnosis of acute pulmonary embolism. Among them, 1,565 (0.8%) had an additional diagnosis of hypocalcemia (Figure 1). The mean age of PE patients with and without hypocalcemia was 61.7 and 62.6 years respectively (p<0.001). There was no significant difference in the racial distribution of patients between the two groups(p=0.062). Patients with hypocalcemia had a higher number of comorbid conditions (Charlson index ≥3-39.9% vs. 28%) than without hypocalcemia. The study did not show any statistical difference in the percentage of males, mean age, race, hospital location, and bed size. The insurance types for patients with and without hypocalcemia was not significantly different (p=0.656) with Medicare being the most common insurance type in both groups (51.8% vs 52%). 12.4% of PE patients with hypocalcemia died in hospital, compared to 2.95% in those without hypocalcemia. On multivariate regression analysis adjusting for confounders, PE and hypocalcemia patients had 4 times higher odds (aOR-4.03, 95% CI 2.78-5.84, p<0.001) of in-hospital mortality compared to those with only PE.

**Figure 1:**
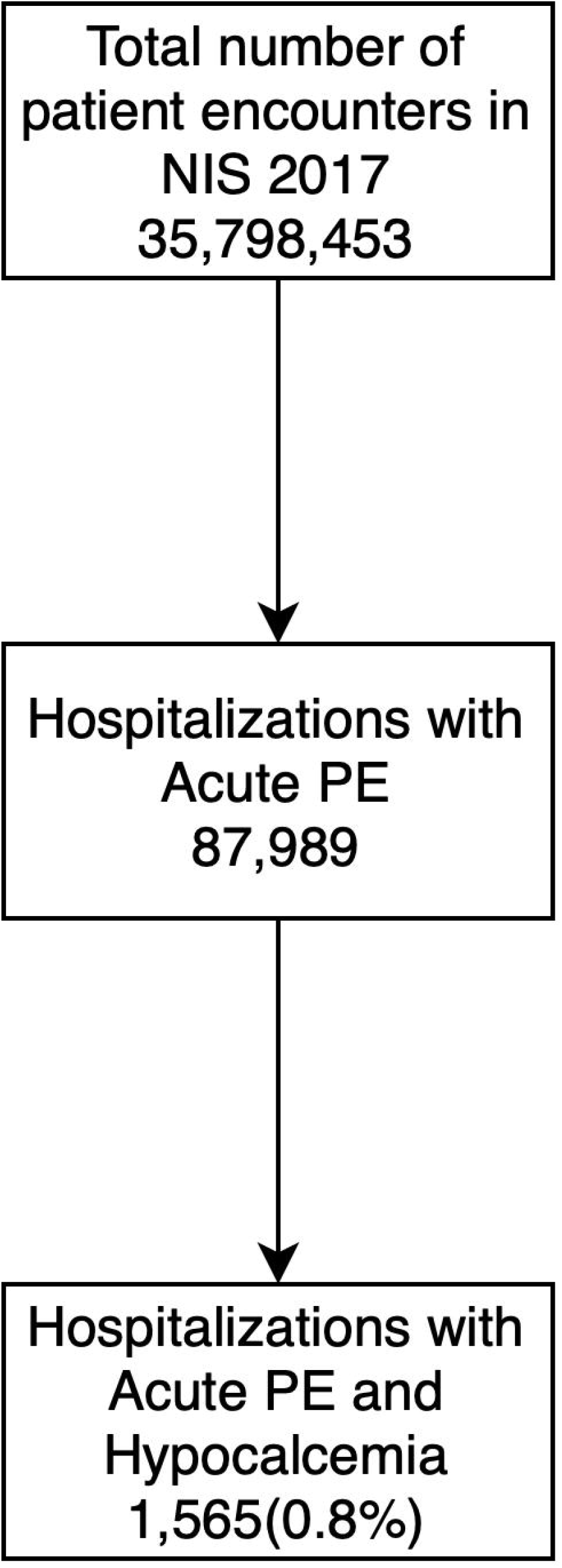
Flowchart showing selection criteria for the patients

On multivariate analysis, the LOS was longer for patients with hypocalcemia (7.41 vs 4.26 days), with an adjusted increase in LOS by 2.8 days (P=<0.001). The incidence of acute kidney injury (aOR=2.62, CI 1.95-3.52 p<0.001), acute respiratory failure (a0R=1.84, CI 1.42-2.38 p<0.001), sepsis (aOR=4.99, CI 3.08-8.11 p<0.001) and arrhythmias (aOR=2.63, CI 1.99-3.48 p<0.001) were also higher for PE patients with hypocalcemia. Using propensity matching patients with Hypocalcemia and PE had 4.4 times higher odds of in-hospital mortality (aOR=4.43, CI 3.03-6.50, p<0.001) compared to those with only PE.

## DISCUSSION

Our study demonstrated worse in-hospital mortality among PE patients with hypocalcemia than without hypocalcemia. The in-hospital complications were also higher, and hypocalcemia patients were hospitalized for 2.8 days longer on average.

Calcium is involved in platelet function and numerous parts of the coagulation cascade((7). Its role in the coagulation cascade may partly explain the association of hypocalcemia with such outcomes. Calcium may also result in arterial relaxation and reduced blood pressure through perivascular receptor activation, leading to hematoma expansion in bleeding patients. Morotti et al. reported such an association in patients with intracerebral hemorrhage (8).

Patients with hypocalcemia had a higher number of comorbid conditions (Charlson index ≥3-39.9% vs 28%). One possible explanation could be the higher prevalence of renal dysfunction in such patients predisposing them to low calcium levels (9). The incidence of acute kidney injury, acute respiratory failure, sepsis, and arrhythmias were higher for PE patients with hypocalcemia. The increased incidence of the above conditions may be partly due to more comorbid conditions in patients with hypocalcemia. Additionally, hypocalcemia itself has been linked to certain complications. Severe hypocalcemia has been linked to life-threatening cardiac arrhythmia and seizures (10). Low calcium levels have been specifically discussed in relation to several neurological and cardiac complications, including seizures, status epilepticus, and coma among other complications, although there is discussion as to whether or not these complications may be explained by other causes unrelated to calcium (10). Moreover, in patients hospitalized with COVID-19 infection, hypocalcemia has been found to be associated with increased risk of acute respiratory failure and mortality, underscoring the risk of more complicated hospitalization courses (11).

Previous studies have identified hypocalcemia as an independent predictor of mortality after acute pulmonary thromboembolism. Reports have indicated that pulmonary thromboembolism patients with hypocalcemia had higher death rates than those without hypocalcemia (5). Additionally, there is an association between hypocalcemia and increased mortality in critically ill patients (12). In critically ill patients, hypocalcemia is thought to occur most often because of heart failure and hyperadrenergic states (13). The mechanism of calcium disequilibrium in critically ill patients may be due to end-organ resistance to parathyroid hormone, impairment of the parathyroid hormone secondary to catecholamine excess or pro-inflammatory cytokine release and inhibited parathyroid hormone release and cellular redistribution of calcium. Vitamin D deficiency or insufficiency may also be present in critically ill patients and is reported to be prevalent in as many as 78% of intensive care unit (ICU) patients (13). Patients with PE may require ICU hospitalizations and may require mechanical ventilation, catheterization, and thrombolytic use (14). These results warrant further studies to confirm these findings and investigate possible mechanisms.

The presence of increased mortality in patients with hypocalcemia may suggest that hypocalcemia may be a useful prognostic tool in PE patients. There are several prognostic assessment tools that exist for PE patients, including the pulmonary embolism severity index (PESI) and the simplified PESI (sPESI) (15, 16). Moreover, imaging and cardiac biomarkers are also used to stratify patients with an intermediate and high risk of PE(17). Previous research has identified hypocalcemia to be an independent predictor of 30-day mortality following a PE. An optimal prediction rule, which contains hypocalcemia and several variables from the PESI and sPESI, has been shown to have higher predictive value and validity than the PESI and sPESI respectively (5). Our findings endorse the relationship between hypocalcemia and in-hospital mortality that has been discussed in the literature, and hypocalcemia may be a valuable future prognostic tool for PE patients.

### Strengths and limitations

One can appreciate several limitations in this study. This study examines numerous demographics of PE hospitalizations, offering a comprehensive and thorough overview of PE hospitalizations. We obtained the population used for this study from the NIS, an extensive, multiethnic hospital-based registry. However, as with any study, there are limitations that should be noted. Data from the NIS is subject to biases associated with retrospective studies. Additionally, the NIS reports information on hospitalizations and not from individual patients; therefore, patients admitted numerous times would be included more than once in the data set. The database also uses ICD-10 codes to report information, leaving it subject to coding errors. Finally, the NIS does not include information about the severity of the diagnosis at the time of admission. For example, it does not include the precise calcium level of patients with hypocalcemia.

Despite the limitations mentioned above, the large sample size, outcomes of the study, and analysis techniques make for a study that provides a comprehensive overview of hypocalcemia and PE while aiming to encourage further discourse and future controlled prospective studies.

## CONCLUSION

Pulmonary embolism (PE) is associated with significant mortality and morbidity, and disorders of calcium metabolism are a commonly encountered medical problem. Our study showed that patients with PE and hypocalcemia had higher in-hospital mortality compared to those without hypocalcemia. These patients also had more comorbid conditions, as reflected by the Charlson comorbidity index. Patients hospitalized with a PE and concurrent hypocalcemia may be at higher risk of numerous in-hospital complications, including sepsis, arrhythmias, acute kidney injury, and death. The findings of our study may reflect the effectiveness of using calcium as a prognostic marker in patients hospitalized with a pulmonary embolism.

## Data Availability

None

## Tables

**Table 1:** Baseline Characteristics of Hospitalizations for PE with and without Hypocalcemia

|  | **PE only, %** | **PE and hypocalcemia, %** | **p-value** |
| --- | --- | --- | --- |
| **Total no.** | 186242 | 1565 |  |
| Mean age (years) | 62.6 | 61.7 | <0.001 |
| Female | 51.9 | 50.8 | 0.67 |
| Race |  |  | 0.062 |
| White | 71.9 | 65.8 |  |
| Black | 18.9 | 22.1 |  |
| Hispanic | 5.5 | 5.9 |  |
| Asians or Pacific Islander | 0.9 | 2.34 |  |
| **Charleston comorbidity index** |  |  | <0.001 |
| 0 | 32.1 | 18.8 |  |
| 1 | 23.3 | 18.8 |  |
| 2 | 16.4 | 22.3 |  |
| ≥3 | 28 | 39.9 |  |
| **Hospital type** |  |  | 0.0009 |
| Rural | 9.8 | 5.7 |  |
| Urban nonteaching | 23.8 | 17.8 |  |
| Urban teaching | 66.3 | 76.3 |  |
| Expected primary payer |  |  | 0.6562 |
| Medicare | 51.8 | 52 |  |
| Medicaid | 12 | 13.7 |  |
| Private | 29.5 | 27.1 |  |
| Self-pay | 3.7 | 3.5 |  |
| **Median household income, in US \$** | | | 0.028 |
| 1 - 43,999 | 28.3 | 33.9 |  |
| 44,000 - 55,999 | 27.2 | 24.9 |  |
| 56,000 - 73,999 | 24.4 | 26.5 |  |
| 74,000+ | 19.9 | 14.5 |  |

**Table 2:** Multivariate Regression Analysis of Primary and Secondary Outcomes

|  | **PE** | **PE and Hypocalcemia** | **Adjusted odds ratio (aOR)** | **P-value** |
| --- | --- | --- | --- | --- |
| **Primary outcome** |  |  |  |  |
| Death | 2.95 | 12.4 | 4.03 (2.78-5.84) | <0.001 |
| **Secondary outcomes** |  |  |  |  |
| AKI | 11 | 24 | 2.62 (1.95-3.52) | <0.001 |
| ARF | 22 | 35 | 1.84 (1.42-2.38) | <0.001 |
| Sepsis | 1.2 | 6.7 | 4.99 (3.08-8.11) | <0.001 |
| Arrhythmias | 16 | 30 | 2.63 (1.99-3.48) | <0.001 |

## Notes

### Competing Interest Statement

The authors have declared no competing interest.

